# Mask mandates can limit COVID spread: Quantitative assessment of month-over-month effectiveness of governmental policies in reducing the number of new COVID-19 cases in 37 US States and the District of Columbia

**DOI:** 10.1101/2020.10.06.20208033

**Authors:** Michael J. Maloney, Nathaniel J. Rhodes, Paul R. Yarnold

## Abstract

**Introduction:** SARS-CoV-2 is the beta-coronavirus responsible for COVID-19. Facemask use has been qualitatively associated with reduced COVID-19 cases, but no study has quantitatively assessed the impact of government mask mandates (*MM*) on new COVID-19 cases across multiple US States.

**Data and Methods:** We utilized a non-parametric machine-learning algorithm to test the *a priori* hypothesis that *MM* were associated with reductions in new COVID-19 cases. Publicly available data were used to analyze new COVID-19 cases from 37 States and the District of Columbia (i.e., “38 States”). We conducted confirmatory All-States and State-Wise analyses, validity analyses [e.g., leave-one-out (LOO) and bootstrap resampling], and covariate analyses.

**Results:** No statistically significant difference in the daily number of new COVID-19 infections was discernable in the All-States analysis. In State-Wise LOO validity analysis, 11 States exhibited reductions in new COVID-19 and the reductions in four of these States (AK, MA, MN, VA) were significant in bootstrap resampling. Only the Social Capital Index predicted *MM* success (training *p*<0.028 and LOO *p*<0.013).

**Conclusion:** Results obtained when studying the impact of *MM* on COVID-19 cases varies as a function of the heterogeneity of the sample being considered, providing clear evidence of Simpson’s Paradox and thus of confounded findings. As such, studies of *MM* effectiveness should be conducted on disaggregated data. Since transmissions occur at the individual rather than at the collective level, additional work is needed to identify optimal social, psychological, environmental, and educational factors which will reduce the spread of SARS-CoV-2 and facilitate *MM* effectiveness across diverse settings.

## INTRODUCTION

The significance of respiratory droplets and airborne viral particles in the spread of SARS-CoV-2 and propagation of the COVID-19 pandemic gained early attention.^1^ Asymptomatic carriage of SARS-CoV-2 is important to community spread because these individuals can transmit the virus in exhaled breath.^2, 3^ Corollary research reported that face masks are protective^4-7^ and may limit the severity of COVID-19 among individuals who become infected.^8^ Indeed, the Centers for Disease Control and Prevention (CDCP) has advocated wearing a facemask when social isolation is impossible or impractical,^9^ and recently reported that dining-out is among the riskiest known activities during the coronavirus pandemic—since face masks *are not used* when people are eating and drinking.^10^ Other activities associated with an increased risk of SARS-CoV-2 transmission include singing^11^ and aerosol generating procedures in the healthcare setting.^12^ Small droplet formation, which can lead to aerosolization of the virus, is a major driver of infection risk,^13^ thus wearing a facemask is expected to reduce inhalation of aerosolized virus.^4^ Indeed, low rates of positive serology were observed (i.e., < 4% n=1/27) when healthcare workers followed aerosol minimizing procedures *and* used personal protective equipment to perform deep respiratory sampling in patients with COVID-19.^14^ Thus, there is consensus in the scientific, clinical, and business communities that appropriate wearing of facemasks is a “best-practice” personal behavior, which can reduce the chances of being infected by, or of infecting others with, SARS-CoV-2 and other airborne disease-causing microbes.

Mathematical and time-series models of COVID-19 data collected in the US investigated salutary effects of masking on reducing the spread of the pandemic.^15, 16^ These models suggest that a consistent application of best-practice public-health orders and citizen behaviors is needed to slow the spread of the pandemic.^17^ A hybrid modeling approach used by the Institute for Health Metrics and Evaluation predicted that if 95% of people in the US wore masks outside their homes, the number of projected deaths from COVID-19 would drop by half within four months.^18^ Thus, experts hypothesize that Mask Mandate (*MM*) orders should result in reduced COVID-19 transmission and fatalities—assuming that the individuals in the community adhere to the *MM*.

Encouraged by research findings that wearing masks reduces the number of new COVID-19 cases, and motivated to intervene by increasing numbers of cases, Governors of 37 US States and the District of Columbia (hereafter called the “38 States”) issued *MM* orders. However, the lack of clear, consistent Global, Federal, and State guidance on the use of face coverings (e.g., mandates *vs*. recommendations), and on who should be using masks (e.g., essential workers *vs*. private citizens), has initiated a nation-wide naturalistic experiment.^15^ In the absence of clear guidance and direct leadership, variations in *MM* acceptance and enforcement have arisen within and between US States.^17, 19-22^

Promulgating and exacerbating confusion regarding the effectiveness of wearing face masks, adjudicating the effectiveness of *MM* orders has largely been left to *qualitative interpretations* and *visual/numerical inspections* of the epidemic curves depicting the number of new cases over time. Qualitative assessment cannot objectively test the *a priori* hypothesis that wearing a mask in public spaces reduces the incidence of COVID-19 cases. Accordingly, using publicly-available data, we employ non-parametric maximum-accuracy machine-learning to *quantitatively* test the confirmatory (*a priori*) hypothesis that imposing a *MM reduced* daily number of new COVID-19 cases in the month *after vs. before* the *MM*.

## METHODS

### Hypotheses

First, merging data from all 38 States in which the Governor issued a *MM*, we test the *a priori* hypothesis that “*All-States*” imposition of a *MM reduced* the daily number of new COVID-19 cases in the month *after vs*. in the month *before* the *MM*. States were included if State leadership (e.g., Governors) had mandated the use of facemasks by all public-facing employees.

Second, considering each State individually, we test the *a priori* hypothesis that “*State-Wise”* imposition of *MM reduced* the daily number of new COVID-19 cases in the month *after vs*. in the month *before* the *MM*.

### Data

The daily number of new COVID-19 cases was obtained separately for each State in the month *before* the *MM* was made (maximum of 30 days), and in the month *after* the *MM* was made (maximum of 31 days). Case reports occurring before the *MM* were dummy-coded as class=0, and case reports occurring after the *MM* were coded as class=1. Data from The New York Times, based on reports from State and local health agencies, were initially downloaded from GitHub on August 8, 2020 (data re-confirmed on September 23, 2020)^23^ and cross referenced against the COVID-19 State Policy Database (data through June 1, 2020) for dates that *MM* were issued in each State.^24^

Our decision to compare daily number of new cases in the month before *vs*. the month after the *MM* was based on methodological and statistical considerations. First, *MM* orders were made because the day-over-day increases in the number of new cases threatened the ability of available health care resources to meet patient and healthcare worker needs. Therefore, the month following the *MM* naturally offers great opportunity for increased mask wearing to meaningfully reduce the number of new cases. Second, some amount of time is required after the *MM* is made for citizens (who wish to comply) to obtain masks, learn how and when to wear them, and how to care for them, and for new infections occurring in the two-week period prior to the *MM* to develop symptoms after the *MM* was made. Third, to maximize the available statistical power of our analyses for estimating short-term effects (see *Statistical Analysis*), we extracted the number of cases for up to 30 days before and up to 31 days after the *MM* was implemented (yielding a maximum of n=61 data points for each state).

Additional attributes, obtained for individual States, were used in an attempt to discriminate States for which a *MM* did *vs*. did not reduce the number of new COVID-19 cases. Numerical attributes included population estimates,^26^ gross domestic product (State and per capita),^27^ age (with arbitrary brackets of 0-18, 18-25, 26-34, 35-54, 55-64, and 65+),^28^ pre-pandemic (January 2020) number of unemployed persons and rate,^29^ homelessness,^30^ shelter beds,^31^ incarceration number and rate,^32^ number of nursing home residents,^33^ population density (people per square mile in 2015),^34^ urban overcrowding (number of houses having >1 person per room),^35^ severe urban overcrowding (number of houses having >1.5 people per room),^35^ ethnicity (white, black, and Hispanic categories),^36^ Social Capital Index (a measure of social cohesion, where higher positive numbers indicate greater societal cohesion),^37^ and obesity.^38^ Categorical attributes included if the State’s Governor wore a mask in public after the *MM* was made (0=Did not wear mask, 1=wore mask) which was assessed using media reports and public database image searches, the Governor’s political party (0=Republican, 1=Democrat),^39^ and the Governor’s gender (0=Female, 1=Male).^40^

### Statistical Analysis

We used confirmatory *<u>O</u>*ptimal *<u>D</u>*iscriminant (or *<u>D</u>*ata) *<u>A</u>*nalysis (ODA) to ascertain the *predictive accuracy* attained by using maximum-accuracy models to evaluate the *hypothesized reduction in the number of new cases* occurring subsequent to the *MM*.^41-43^ ODA is the moniker of the statistically-motivated non-parametric machine-learning algorithm which identifies the cut-point for an ordered *attribute* (i.e., “independent variable”), or the assignment rule for a categorical attribute, which yields the *strongest achievable accuracy* in discriminating between two or more class categories (i.e., “dependent variable”) for a given sample and hypothesis.^41-43^ In the present study all hypotheses were confirmatory.^42^ For both the All-States (38-State) and the State-Wise analysis, the *a priori* hypothesis was that the *MM* (up to 31 days) reduced the number of new cases.^44, 45^

An ODA model is obtained by iterating through every possible different assignment rule that is consistent with the *a priori* hypothesis, and identifying the model achieving the highest effect strength for sensitivity (ESS) statistic.^41, 42^ An index of classification accuracy adjusted to remove the effect of chance, ESS is a function of the mean sensitivity of the model achieved across class categories, standardized such that 0 represents the discriminatory accuracy which is expected by chance, 100 represents errorless (perfect) discrimination, and values of −100 ≤ ESS < 0 represent accuracy which is worse than expected by chance. For a two-category *class variable* such as studied herein (i.e., month before *MM*=class 0 *vs*. month after *MM*=class 1),

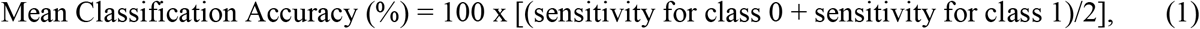

and

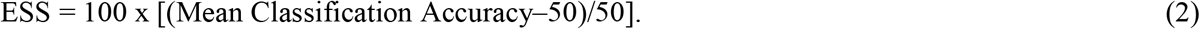

A nonparametric permutation test which requires no distributional assumptions is used to assess the statistical significance (*p* value) of the achieved ESS.^41, 44, 45^ Point estimates of *p* values for tests of statistical hypotheses reported herein are indicated as being statistically significant (*p*≤0.05) at either the *experimentwise* (confirmatory *p*≤*p*_Bonferroni_) or the *per-comparison* (confirmatory *p*≤0.05) criterion.^42^

Internal model validation was conducted using bootstrap resampling^46, 47^ (n=25,000) of the State-Wise models within the *ODA* package for R.^48^ Specifically, resampling with 50% replacement was conducted using the LOO (i.e., validity) ODA model confusion matrix for each State. Exact discrete 95% confidence intervals (CI) for the given ODA model (“Model”) and for randomly scrambled observations from the model (“Chance”) were obtained.^46^ Graphical depictions of new cases organized across days, and histograms of bootstrapped 95% CIs, were created using *ggplot2* for R.^49^

Potential generalizability of the ODA model applied to classify an independent random sample is assessed using a one-sample “leave-one-out” (LOO) jackknife analysis.^42^ The sample size offers greater than 90% power to identify an ODA model of moderate strength (i.e., 25≤ESS<50).^50^

Exploratory ODA analyses were conducted to model the association of inter-State differences 28 variables (e.g., age, wealth, population density) conceivably capable of discriminating States which did *vs*. did not exhibit the predicted decline in number of new cases in the month following a *MM* (see *Data Supplement*).^42^

Sensitivity analyses were conducted comparing the initial data (obtained August 8, 2020) to the data obtained September 23, 2020. The percentage difference between the two datasets were calculated for each value [i.e., 100 x (data_new_-data_old_) / data_old_] to quantify differences between datasets. In sensitivity analyses, any differences between datasets were compared according to the ODA model cut-points obtained using the initial data to determine whether the model was invariant to new (updated) data.

## RESULTS

### All-States Analysis

The *mean* number of new COVID-19 cases before *vs*. after the *MM* for the sample of 38 States was 654 (N=1138, SD=1357) *vs*. 639 (N=1177, SD=975), respectively. In three instances, the daily report of new cases appeared to be outliers (GA 4/12/20, LA 5/29/20, and VA 5/6/20) and were coded as missing (i.e., coded as −999). Confirmatory ODA was used to test the *a priori* hypothesis that imposition of a *MM* reduced the *number* of new COVID-19 cases (the attribute) one-month *after vs*. one-month *before* the *MM* (the class variable). The confirmatory ODA model was: if >4 new cases-per-day then predict pre-*MM*; otherwise if ≤4 new cases-per-day, then predict post-*MM*. In training (using all observations from the sample) analysis, this model correctly classified 98.1% of the pre-*MM* cases, and 6.1% of the post-*MM* cases: ESS=4.1 (a weak effect), *p*<0.13.

Thus, when considering all 38 States having a *MM* as a single sample, there was no statistically significant difference in the daily number of new COVID-19 infections in the month before *vs*. the month after the *MM*.

### State-Wise Analysis

As seen in Table 1, after the *MM* the *mean* number of new daily cases was numerically *lower* in 13 States (AK, GA, HI, LA, MA, MI, MN, NY, VA, VT, WA, WV, WY) post-*MM*, and numerically *higher* in the remaining 25 States. Table 2 gives the findings of ODA used to test the *a priori* hypothesis that State-Wise imposition of a *MM* (made by 38 different State Governors) *reduced the number of new COVID-19 cases* in the month *after vs*. in the month *before* the *MM*—with each State considered individually. In each State-Wise analysis, the class variable was a binary coded variable signifying the period before *vs*. after the *MM* occurred, and the attribute was the daily number of new cases. In Table 2, the States are ordered from strongest to weakest ESS of the confirmatory ODA model in training analysis.

**Table 1:** State-Wise Descriptive Statistics for Number of New Cases Pre- *vs*. Post-Mask Mandate (*MM*) SD=Standard Deviation

| State | <u>Before *MM*</u><br>(n≤30) |  | <u>After *MM*</u><br>(n≤31) |  |
| --- | --- | --- | --- | --- |
|  | Mean | SD | Mean | SD |
| AK | 9.8 | 4.8 | 2.4 | 2.0 |
| AL | 229.4 | 64.1 | 390.3 | 158.0 |
| AR | 93.7 | 75.4 | 205.0 | 114.8 |
| AZ | 245.6 | 88.2 | 547.7 | 355.1 |
| CA | 1417.9 | 390.6 | 2147.9 | 464.8 |
| CO | 338.6 | 81.2 | 421.1 | 148.5 |
| DC | 68.0 | 52.3 | 155.3 | 52.7 |
| DE | 147.2 | 100.1 | 153.7 | 69.2 |
| FL | 754.3 | 263.8 | 863.7 | 305.2 |
| GA | 704.1 | 315.6 | 653.4 | 218.1 |
| HI | 17.1 | 9.4 | 3.4 | 4.3 |
| IL | 1564.2 | 504.7 | 2182.9 | 668.9 |
| IN | 530.7 | 134.4 | 559.0 | 132.2 |
| KY | 159.5 | 87.9 | 178.8 | 85.7 |
| LA | 760.2 | 606.1 | 399.2 | 237.0 |
| MA | 1925.7 | 718.4 | 1041.5 | 660.5 |
| MD | 385.2 | 292.1 | 911.0 | 240.9 |
| ME | 26.4 | 12.6 | 39.7 | 16.7 |
| MI | 1147.5 | 344.6 | 572.2 | 255.6 |
| MN | 637.7 | 116.4 | 383.6 | 101.5 |
| MS | 222.9 | 68.5 | 277.7 | 81.8 |
| NE | 179.6 | 152.3 | 296.1 | 109.6 |
| NH | 59.3 | 17.4 | 80.8 | 29.6 |
| NJ | 1480.3 | 1528.0 | 2936.7 | 808.8 |
| NM | 117.1 | 47.0 | 146.3 | 51.3 |
| NV | 120.8 | 38.8 | 124.9 | 42.7 |
| NY | 7479.5 | 2836.8 | 4170.2 | 2206.4 |
| OH | 503.5 | 288.7 | 574.1 | 106.9 |
| OR | 61.0 | 14.9 | 59.9 | 29.0 |
| PA | 1051.8 | 605.8 | 1150.4 | 355.1 |
| RI | 138.1 | 120.7 | 278.0 | 87.1 |
| TX | 923.8 | 222.6 | 1266.6 | 380.9 |
| UT | 65.9 | 56.0 | 137.8 | 34.7 |
| VA | 933.2 | 243.9 | 656.0 | 230.7 |
| VT | 25.1 | 16.2 | 5.5 | 5.2 |
| WA | 299.2 | 132.8 | 252.7 | 108.5 |
| WV | 31.8 | 17.7 | 28.4 | 19.5 |
| WY | 14.6 | 12.9 | 11.1 | 6.6 |

**Table 2:**
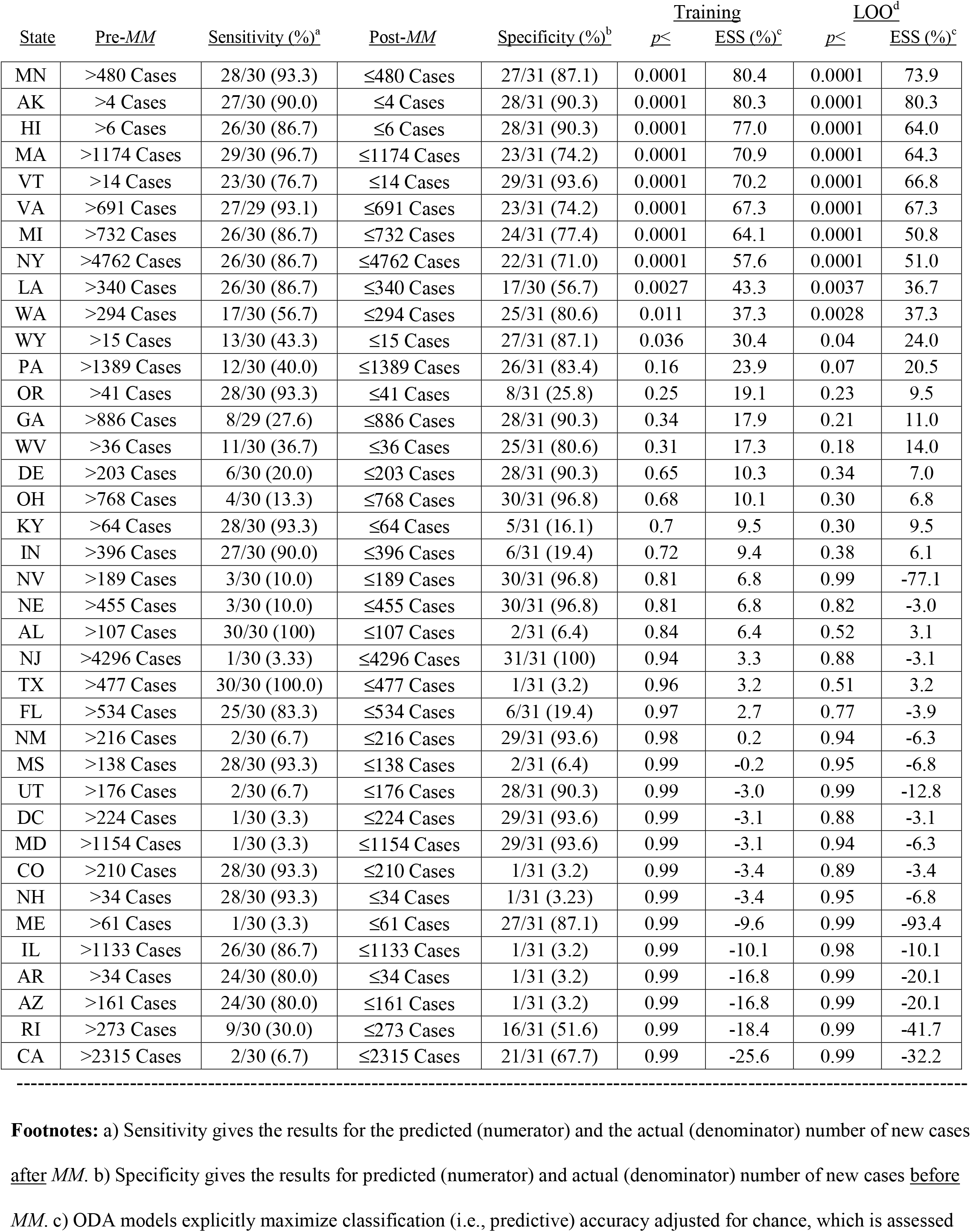
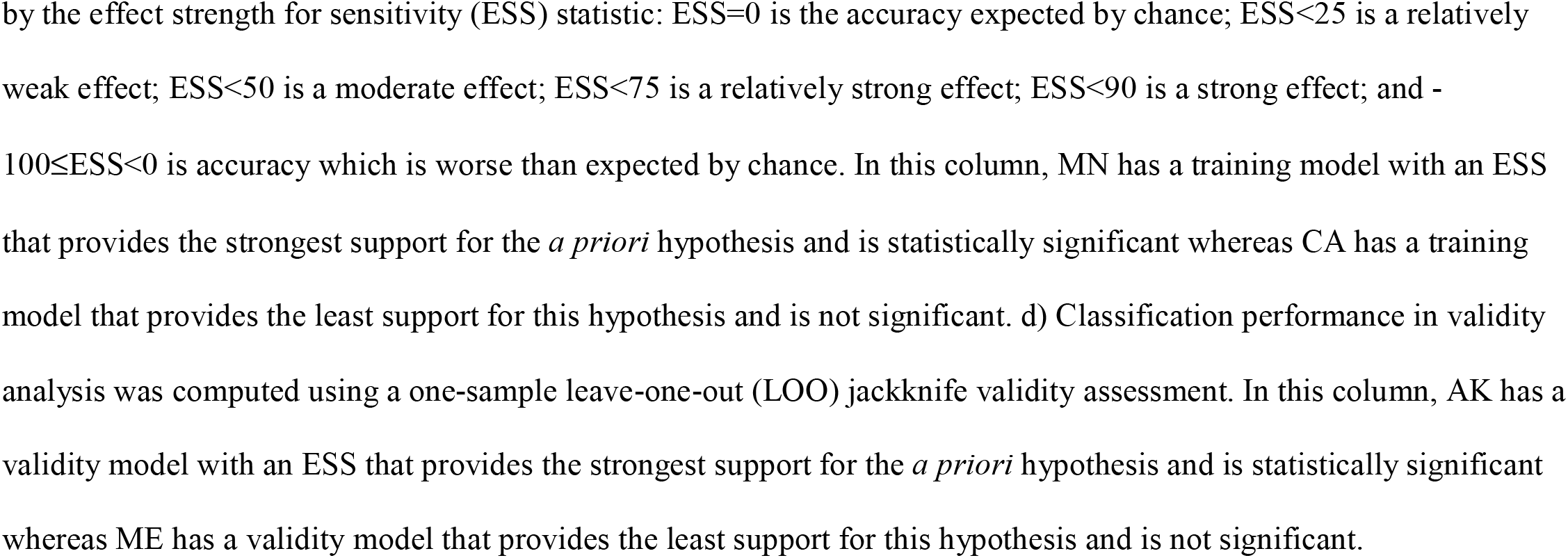
Maximum-Accuracy Confirmatory Models Assessing State-Wise Mask Mandate (*MM*) Effectiveness

| State | Pre-*MM* | Sensitivity (%)<sup>a</sup> | Post-*MM* | Specificity (%)<sup>b</sup> | Training |  | LOO<sup>d</sup> |  |
| --- | --- | --- | --- | --- | --- | --- | --- | --- |
|  |  |  |  |  | *p*≤ | ESS (%)<sup>c</sup> | *p*≤ | ESS (%)<sup>c</sup> |
| MN | >480 Cases | 28/30 (93.3) | ≤480 Cases | 27/31 (87.1) | 0.0001 | 80.4 | 0.0001 | 73.9 |
| AK | >4 Cases | 27/30 (90.0) | ≤4 Cases | 28/31 (90.3) | 0.0001 | 80.3 | 0.0001 | 80.3 |
| HI | >6 Cases | 26/30 (86.7) | ≤6 Cases | 28/31 (90.3) | 0.0001 | 77.0 | 0.0001 | 64.0 |
| MA | >1174 Cases | 29/30 (96.7) | ≤1174 Cases | 23/31 (74.2) | 0.0001 | 70.9 | 0.0001 | 64.3 |
| VT | >14 Cases | 23/30 (76.7) | ≤14 Cases | 29/31 (93.6) | 0.0001 | 70.2 | 0.0001 | 66.8 |
| VA | >691 Cases | 27/29 (93.1) | ≤691 Cases | 23/31 (74.2) | 0.0001 | 67.3 | 0.0001 | 67.3 |
| MI | >732 Cases | 26/30 (86.7) | ≤732 Cases | 24/31 (77.4) | 0.0001 | 64.1 | 0.0001 | 50.8 |
| NY | >4762 Cases | 26/30 (86.7) | ≤4762 Cases | 22/31 (71.0) | 0.0001 | 57.6 | 0.0001 | 51.0 |
| LA | >340 Cases | 26/30 (86.7) | ≤340 Cases | 17/30 (56.7) | 0.0027 | 43.3 | 0.0037 | 36.7 |
| WA | >294 Cases | 17/30 (56.7) | ≤294 Cases | 25/31 (80.6) | 0.011 | 37.3 | 0.0028 | 37.3 |
| WY | >15 Cases | 13/30 (43.3) | ≤15 Cases | 27/31 (87.1) | 0.036 | 30.4 | 0.04 | 24.0 |
| PA | >1389 Cases | 12/30 (40.0) | ≤1389 Cases | 26/31 (83.4) | 0.16 | 23.9 | 0.07 | 20.5 |
| OR | >41 Cases | 28/30 (93.3) | ≤41 Cases | 8/31 (25.8) | 0.25 | 19.1 | 0.23 | 9.5 |
| GA | >886 Cases | 8/29 (27.6) | ≤886 Cases | 28/31 (90.3) | 0.34 | 17.9 | 0.21 | 11.0 |
| WV | >36 Cases | 11/30 (36.7) | ≤36 Cases | 25/31 (80.6) | 0.31 | 17.3 | 0.18 | 14.0 |
| DE | >203 Cases | 6/30 (20.0) | ≤203 Cases | 28/31 (90.3) | 0.65 | 10.3 | 0.34 | 7.0 |
| OH | >768 Cases | 4/30 (13.3) | ≤768 Cases | 30/31 (96.8) | 0.68 | 10.1 | 0.30 | 6.8 |
| KY | >64 Cases | 28/30 (93.3) | ≤64 Cases | 5/31 (16.1) | 0.7 | 9.5 | 0.30 | 9.5 |
| IN | >396 Cases | 27/30 (90.0) | ≤396 Cases | 6/31 (19.4) | 0.72 | 9.4 | 0.38 | 6.1 |
| NV | >189 Cases | 3/30 (10.0) | ≤189 Cases | 30/31 (96.8) | 0.81 | 6.8 | 0.99 | -77.1 |
| NE | >455 Cases | 3/30 (10.0) | ≤455 Cases | 30/31 (96.8) | 0.81 | 6.8 | 0.82 | -3.0 |
| AL | >107 Cases | 30/30 (100) | ≤107 Cases | 2/31 (6.4) | 0.84 | 6.4 | 0.52 | 3.1 |
| NJ | >4296 Cases | 1/30 (3.33) | ≤4296 Cases | 31/31 (100) | 0.94 | 3.3 | 0.88 | -3.1 |
| TX | >477 Cases | 30/30 (100.0) | ≤477 Cases | 1/31 (3.2) | 0.96 | 3.2 | 0.51 | 3.2 |
| FL | >534 Cases | 25/30 (83.3) | ≤534 Cases | 6/31 (19.4) | 0.97 | 2.7 | 0.77 | -3.9 |
| NM | >216 Cases | 2/30 (6.7) | ≤216 Cases | 29/31 (93.6) | 0.98 | 0.2 | 0.94 | -6.3 |
| MS | >138 Cases | 28/30 (93.3) | ≤138 Cases | 2/31 (6.4) | 0.99 | -0.2 | 0.95 | -6.8 |
| UT | >176 Cases | 2/30 (6.7) | ≤176 Cases | 28/31 (90.3) | 0.99 | -3.0 | 0.99 | -12.8 |
| DC | >224 Cases | 1/30 (3.3) | ≤224 Cases | 29/31 (93.6) | 0.99 | -3.1 | 0.88 | -3.1 |
| MD | >1154 Cases | 1/30 (3.3) | ≤1154 Cases | 29/31 (93.6) | 0.99 | -3.1 | 0.94 | -6.3 |
| CO | >210 Cases | 28/30 (93.3) | ≤210 Cases | 1/31 (3.2) | 0.99 | -3.4 | 0.89 | -3.4 |
| NH | >34 Cases | 28/30 (93.3) | ≤34 Cases | 1/31 (3.23) | 0.99 | -3.4 | 0.95 | -6.8 |
| ME | >61 Cases | 1/30 (3.3) | ≤61 Cases | 27/31 (87.1) | 0.99 | -9.6 | 0.99 | -93.4 |
| IL | >1133 Cases | 26/30 (86.7) | ≤1133 Cases | 1/31 (3.2) | 0.99 | -10.1 | 0.98 | -10.1 |
| AR | >34 Cases | 24/30 (80.0) | ≤34 Cases | 1/31 (3.2) | 0.99 | -16.8 | 0.99 | -20.1 |
| AZ | >161 Cases | 24/30 (80.0) | ≤161 Cases | 1/31 (3.2) | 0.99 | -16.8 | 0.99 | -20.1 |
| RI | >273 Cases | 9/30 (30.0) | ≤273 Cases | 16/31 (51.6) | 0.99 | -18.4 | 0.99 | -41.7 |
| CA | >2315 Cases | 2/30 (6.7) | ≤2315 Cases | 21/31 (67.7) | 0.99 | -25.6 | 0.99 | -32.2 |

**Footnotes:** a) Sensitivity gives the results for the predicted (numerator) and the actual (denominator) number of new cases after *MM*. b) Specificity gives the results for predicted (numerator) and actual (denominator) number of new cases before *MM*. c) ODA models explicitly maximize classification (i.e., predictive) accuracy adjusted for chance, which is assessed
by the effect strength for sensitivity (ESS) statistic: $ESS=0$ is the accuracy expected by chance; $ESS<25$ is a relatively weak effect; $ESS<50$ is a moderate effect; $ESS<75$ is a relatively strong effect; $ESS<90$ is a strong effect; and $100\leq ESS<0$ is accuracy which is worse than expected by chance. In this column, MN has a training model with an ESS that provides the strongest support for the *a priori* hypothesis and is statistically significant whereas CA has a training model that provides the least support for this hypothesis and is not significant. d) Classification performance in validity analysis was computed using a one-sample leave-one-out (LOO) jackknife validity assessment. In this column, AK has a validity model with an ESS that provides the strongest support for the *a priori* hypothesis and is statistically significant whereas ME has a validity model that provides the least support for this hypothesis and is not significant.

Among all 38 States, training data for Minnesota (MN, the first State in Table 2) offer the greatest support for the *a priori* hypothesis. The ODA model is: if >480 new cases-per-day, then predict pre-*MM*; otherwise if ≤480 cases-per-day, then predict post-*MM*. This ODA model correctly classified 28 of 30 (93.3%) of the pre-*MM* days, and 27 of 31 (87.1%) of the post-*MM* days, thereby yielding a *strong*^42^ ESS=80.4, *p*<0.0001. However, this model showed modest instability in LOO jackknife analysis (*relatively strong*^42^ ESS=73.9, *p*<0.0001). In contrast, data for Alaska (AK, second row in Table 2) had the second-strongest training effect (*strong* ESS=80.3, *p*<0.0001), which was stable in LOO analysis. Among all 38 States, only AK returned a *strong* confirmation of the *a priori* hypothesis in training and LOO analysis.

Of the total of 38 States included in this study, 11 (23.7%) achieved confirmatory *p*<0.05 at the experimentwise criterion, and two (5.3%) achieved confirmatory *p*<0.05 at the per-comparison criterion^42^ (Table 2). Three States (MN, AK, HI) yielded a training model indicating a *strong* confirmation^42^ (i.e., ESS≥75) of the *a priori* hypothesis. Five States (MA, VT, VA, MI, NY) yielded a training model indicating a *relatively strong* confirmation^42^ (i.e., 50≤ESS<75). And, three States (LA, WA, WY) yielded a training model indicating a *moderate* confirmation^42^ (i.e., 25≤ESS<50) of the *a priori* hypothesis.

Except for GA and WY, States having a numerically greater *mean* number of new daily cases pre- *vs*. post-*MM* (Table 1) also had statistically-significant (at either the experimentwise or the per-comparison criterion^42^) confirmatory (Table 2) and LOO (Table 3) ODA models supporting the *a priori* hypothesis.

**Table 3:**
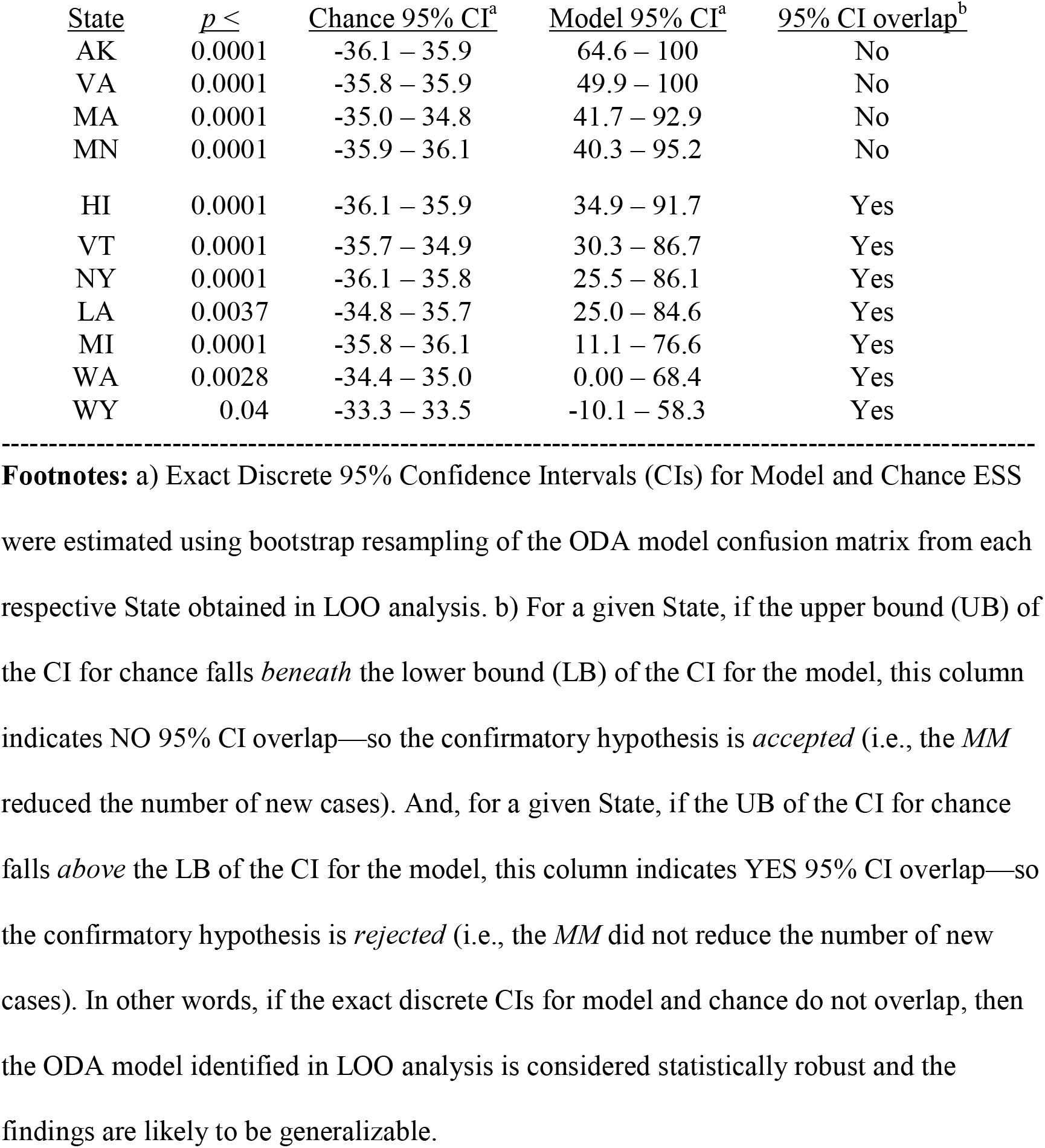
LOO Confirmatory *p*, and Bootstrap 95% CI for LOO ESS, for 11 States Demonstrating Mask Mandate (*MM*) Effectiveness vis-à-vis Training *p*<0.05

For all 11 States in Table 2 having training *p*<0.05, Table 3 presents 95% exact discrete CIs obtained for model and for chance.^46^ For AK, VA, MA, and MN, the *lower bound* of the 95% exact discrete CI *for model* is greater than the *upper bound* of the 95% exact discrete CI *for chance*—therefore the result is statistically significant. For the remaining seven States, the upper bound for chance exceeded the lower bound for the model, so the result was not statistically significant in bootstrap validity analysis.

For example, the daily number of new COVID-19 cases one-month pre- and one-month post-*MM* is illustrated in Figure 1.A for MN. Figure 1.B is a graphical summary of the model and chance bootstrap results: the blue histogram represents chance, the red histogram represents the model, the Y axis is frequency of the result in the Bootstrap analysis, and the X axis is training ESS. The dashed vertical lines inside the Figure represent the lower 95% CI for the model (red dashes), and the upper 95% CI for chance (blue dashes). When the blue dashes are on the same side as the blue histogram, and the red dashes are on the same side as the red histogram, the effect is statistically significant. Thus, evaluating MN in training analysis, the *a priori* hypothesis is accepted with experimentwise *p*<0.05.

**Figure 1.**
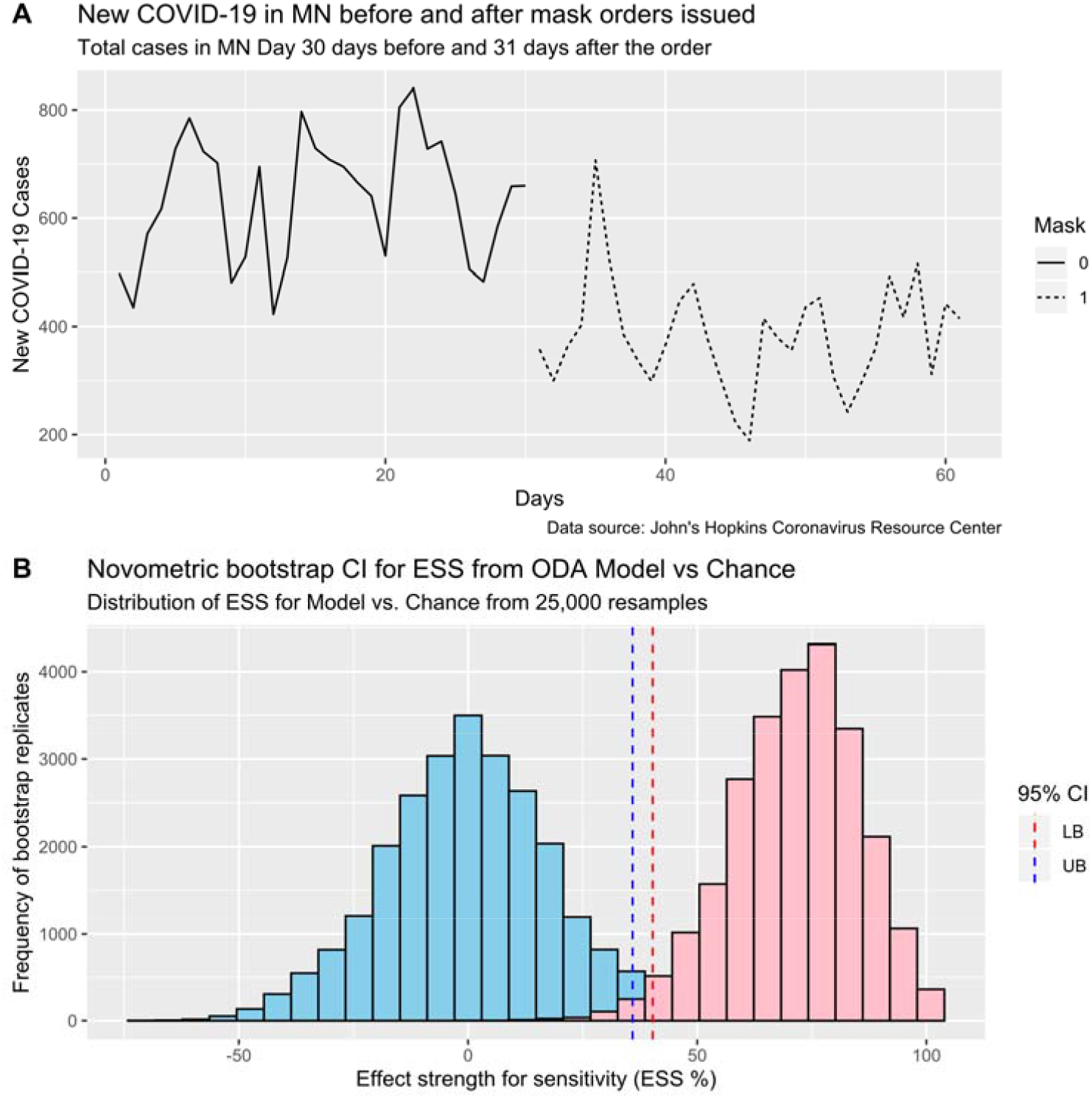
Representative state (MN) with a strong impact of Mask Mandate (*MM*) orders Panel A. Number of new COVID-19 cases reported within MN daily in the month before (solid black line) *vs*. daily in the month after (broken black line) *MM* orders were issued. Panel B. Distribution of bootstrap replicates for model and chance for the LOO analysis for MN by ODA. Here, the ODA model was resampled using bootstrap resampling with replacement (n=25,000 resamples with 50% replacement) for the LOO model confusion matrix (see Table 3) as well as for chance (i.e., the final model predictions randomly scrambled relative to the class variable). The lack of overlap between the Chance UB and Model LB is denoted by the alignment (rather than reversal) of the dashed lines with the corresponding Chance (blue) and Model (red) bootstrap distributions. **Abbreviations**: LB = lower bound of the 95% CI (i.e., 2.5^th^ percentile); UB = upper bound of the 95% CI (i.e., 97.5^th^ percentile); LOO = leave-one-out one-sample jackknife.^42^

In contrast, Figure 2.A presents the daily number of new COVID-19 cases one-month pre- and one-month post-*MM* for WA, and Figure 2.B is a graphical summary of the model and chance bootstrap results. Since the blue dashes are on the side of the red histogram, and the red dashes are on the side of the blue histogram, the hypothesized effect is not statistically significant for WA.

**Figure 2.**
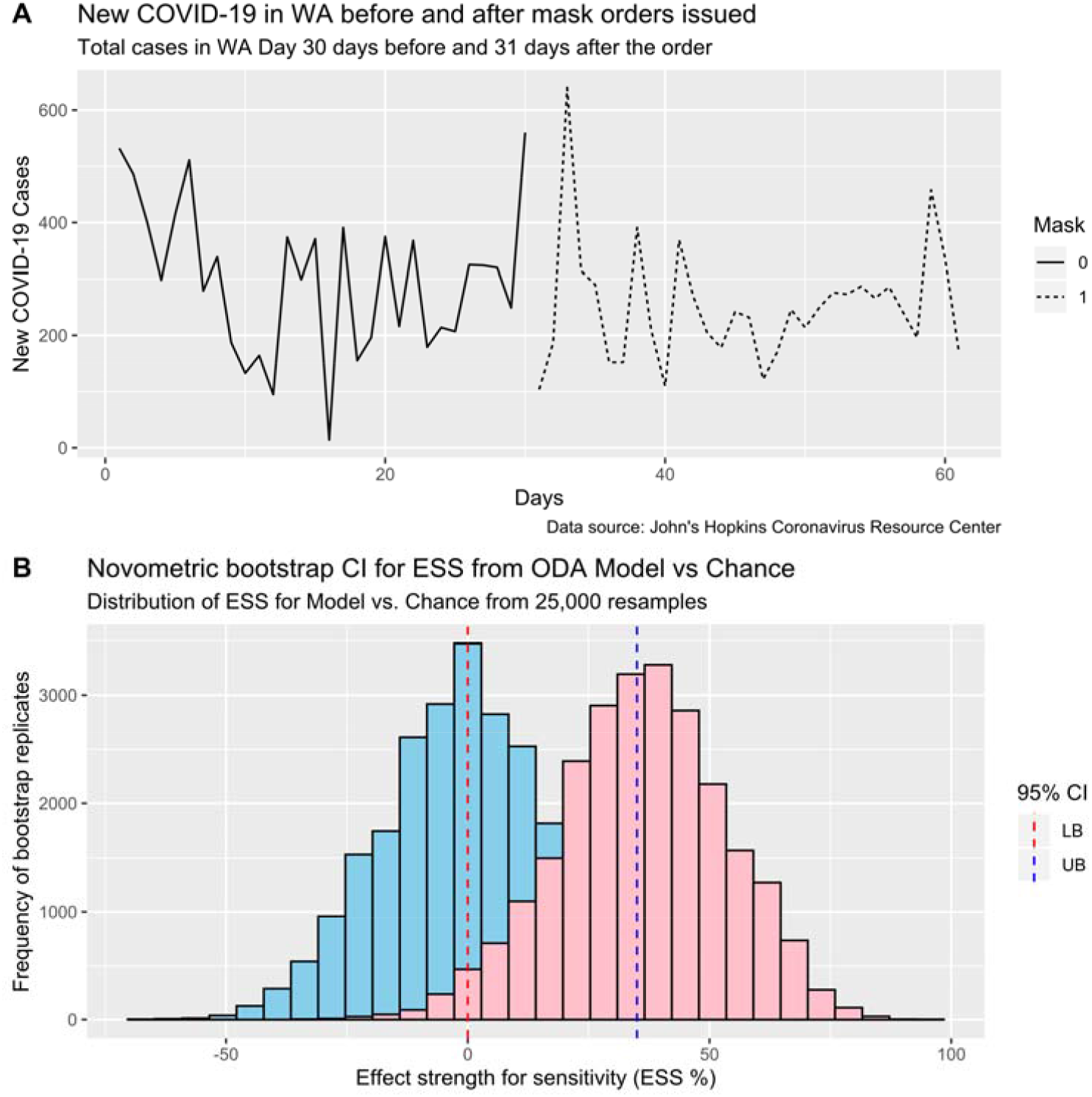
Representative state (WA) with a moderate impact of Mask Mandate (*MM*) orders Panel A. Number of new COVID-19 cases reported within WA daily in the month before (solid black line) *vs*. daily in the month after (broken black line) *MM* orders were issued. Panel B. Distribution of bootstrap replicates for model and chance for the LOO analysis for WA by ODA. Here, the ODA model was resampled using bootstrap resampling with replacement (n=25,000 resamples with 50% replacement) for the LOO model confusion matrix (see Table 3) as well as for chance (i.e., the final model predictions randomly scrambled relative to the class variable). The overlap between the Chance UB and Model LB is denoted by the reversal (rather than alignment) of the dashed lines with the corresponding Chance (blue) and Model (red) bootstrap distributions. **Abbreviations**: LB = lower bound of the 95% CI (i.e., 2.5^th^ percentile); UB = upper bound of the 95% CI (i.e., 97.5^th^ percentile); LOO = leave-one-out one-sample jackknife.^42^

### Modeling Success *vs*. Failure of a *MM* Order in Reducing the Number of New COVID-19 Cases

Finally, ODA was used to discriminate the four States (AK, MA, MN, VA) having a significantly lower number of new cases after the *MM vs*. the 34 States which did not support the *a priori* hypothesis. The only attribute which met the per-comparison criterion for statistical significance^42^ was SCI, a measure of societal cohesion and the closeness of social ties.^51^ The ODA model was: if SCI≤0.311, then predict the *MM did not reduce* the number of new cases of COVID-19; otherwise if SCI>0.311 then predict the *MM reduced* the number of new cases. In training (*p*<0.028) and LOO (*p*<0.013) analysis this model correctly classified all four States in which the *MM* reduced the number of new cases, and 25 of 35 (71.43%) States in which the *MM* failed to reduce the number of new cases. This level of classification accuracy corresponds to ESS=71.43, indicating a relatively strong effect.^42^

### Sensitivity analyses

The dataset obtained on August 8, 2020 to was compared to a dataset obtained on September 23, 2020 and after exclusion of outliers (n=3), case numbers in the new dataset matched the original dataset (i.e., 0% difference) for all states except Oregon (n=12/61) and Virginia (n=5/12), for which the new data fell within ±6% of the original data. These numerical changes in data values (none of which were near the ODA model cut-points) did not change the ODA model, the accuracy (ESS), or statistical significance (*p*) of the model in training or LOO analysis for any model.

## DISCUSSION

An *All-States* analysis was conducted using merged data for 38 US States. Results indicated *no statistically significant decrease* in the number of new daily COVID-19 cases in the month before *vs*. after the *MM* was made. In contradistinction, a *State-Wise* analysis identified *clear evidence of State-specific MM effectiveness in reducing the number of new cases within one month of enactment* for multiple States (Tables 2 and 3). Such clear inconsistency is attributable to improper analysis of merged samples creating an analytic anomaly called *Simpson’s Paradox*—which may identify spurious relationships, may fail to identify actual relationships, and/or may over- or under-weight the strength of effects—in every area of quantitative empirical science.^42, 47^ *Paradoxical confounding* exists when the finding obtained by statistical analysis conducted for a combined sample is different than the finding obtained by the same analysis conducted separately for the constituent samples.^52^ In the setting of a public health emergency, such as the current pandemic, accurate knowledge of the effectiveness of public health interventions is essential so that public health authorities can communicate this to the public.

Combining disparate groups is the primary operating procedure in government, scientific, and media discussions about the number of new and cumulative daily, monthly, and total number of COVID-19 cases and fatalities. Examples of such combined groups include global (the maximum-possible combined group), regional (e.g., the northern hemisphere), continental (e.g., Europe, Africa), and country (e.g., China, US, India, Brazil, France, UK, Israel) daily and cumulative case counts.^53^ However, disparities in regional increases in numbers of new cases prompted some analysts to issue a warning against focusing on National trends.^54^ Findings presented herein—that four of 38 (10.5%) States reliably demonstrated the hypothesized reduction in number of new cases in the month after the *MM* was made—similarly suggest a warning against focusing on State trends. Cognizance of the need to combine groups in a manner that prevents invalid statistical conclusions begs the question of how small must a “catchment area” be to count new cases so as to prohibit paradoxical confounding. Unfortunately, it has been shown that Simpson’s Paradox can even occur for single-subject, “N-of-1” designs.^55^ Thus, while accurate case counts and definitions are important for tracking the epidemiology of the pandemic, such measures do not necessarily reflect the “reality on the ground” in any specific community, nor can they reliably discern the individual effectiveness of public health interventions.

The current methodology of conducting after-the-fact pandemic accounting for convenient and familiar (statistically unmotivated) groups is obviously inadequate when it is considered from a problem-solving perspective that focuses on getting (and staying) ahead of the pandemic curve. Others have argued, early in the pandemic, that rapid and comprehensive contact tracing will empower understanding of the situations and the processes underlying new COVID-19 infections.^56^ As is the case in fighting a wildfire, progress is made by going to the active location each day, extinguishing blazes, and reducing available fuel, rather than by assessing the number of acres which burned the day before. However, contact tracing is most effective when community spread is low, so multifaceted interventions are needed to control the spread of the pandemic and presently wearing a mask is the most efficacious known means of slowing the pandemic.^5, 6, 8, 15, 16^

Temporal counts of the number of new cases are an imprecise measure—including the month-over-month methodology used presently—for multiple reasons. Even with perfect *MM* compliance, there is an up to 14-day lag time between infection and presentation of symptoms, and potentially an even longer lag between infection and presentation at a healthcare facility. After a *MM* order is made, there are logistical problems in getting masks to people, and there is a learning curve for users—where, when and how to wear and care for masks. *MM* compliance is imperfect, and enforcement is difficult. Indeed, individual *MM* compliance is expected to be heterogeneous based on experimental evidence from behavioral psychology. Individuals are susceptible to cognitive biases and often use heuristics to address complex problems when uncertainty is high. Examples of such biases and heuristics related to *MM* use include availability bias (e.g., knowing someone who is sick *vs*. not), anchoring bias (e.g., fixation on initial recommendations to avoid mask use to preserve PPE), and substitution heuristics (e.g., physical discomfort with wearing a mask is substituted for mask efficacy).^57^ The impact of these and other biases and heuristics on mask effectiveness is currently understudied.

Because transmissions occur at the level of the individual, rather than at a State or National level, governments should emphasize best-practice personal behaviors, adequate contact tracing, and minimization of high-risk exposures. Indeed, we posit that the efficacy of mask wearing is not at question as this depends only on the use of the mask. Instead, the effectiveness of *MM* can be eroded by multiple factors including asymptomatic carriage, willful ignorance of *MM* and lack of knowledge of how and when to wear masks. Moreover, our State-Wise analysis supports the *a priori* hypothesis that *MM* do indeed have a significant salutary effect on the spread of the pandemic, but more work is needed to define the social, psychological, environmental, and educational facilitators that maximize *MM* effectiveness.

Experimentally speaking, the best test of public health intervention effectiveness (e.g., *MM*)—using a methodology called an “A (before intervention), B (after intervention)” design—would involve simultaneous uniform application, enforcement, and education around the intervention, coupled with exacting real-time contact-tracing. A science-based, consistent approach to disease management could reduce public misunderstanding in this realm, and diminish mistrust of the other public health interventions (e.g., vaccines currently under development).^58^ Our analysis is limited because we were not able to utilize a rigorous AB analytic framework to test our *a priori* hypotheses. Rather, we took advantage of the naturalistic experiment created by the heterogeneous response to the pandemic across States to highlight the paradoxical results obtained when applying an All-States followed by a State-Wise analytic approach. Our analysis was limited to testing our *a priori* hypothesis and it is worth considering the time-dependent impact of *MM*. Additional work is needed to evaluate the impact of *MM* on the growth of the pandemic. Our analysis is conservative in that we focused only on reductions in new cases and may underestimate the true impact of *MM*. Future studies should evaluate the impact of MM on new deaths. Large-scale serological studies may provide more insights into the spread of SARS-CoV-2 *MM* in States with vs. those without early *MM*.

Finally, we found that SCI, a measure of social closeness and ties,^51^ was a relatively strong predictor of *MM* effectiveness. This finding is notable in that SCI could be a facilitating factor that makes mandated public health interventions more efficient. For example, populations with higher levels of social cohesion are inherently more likely to work together when faced with a common problem based on a shared sense of identity and social bonds. However, this finding has limitations. States with higher SCI should not be interpreted as being inherently better at reducing new SARS-CoV-2 cases, but rather we believe this finding also supports a new corollary hypothesis that States with lower SCI may require different and multifaceted interventions to address the pandemic.

## CONCLUSION

We identified paradoxical confounding whereby the effect of *MM* orders when analyzed using the pooled data from all 38 States available differed when compared to a State-Wise analysis. Overemphasis placed on National or State-level trends may obscure the true effectiveness of important public health interventions such as mask wearing, because transmission events are stochastic and occur at the level of individuals. Thus, greater emphasis on the impact of personal behaviors on *MM* effectiveness and thorough contact tracing are needed to combat the pandemic. While societal factors reflected by measures like the SCI may facilitate the efficiency of interventions like *MM* in select regions, more work is needed to define the optimal set of interventions that reduce SARS-CoV-2 transmission for socially diverse populations. Finally, optimal experimental designs, measurements, and analytic approaches are needed to provide the public with a clearer understanding of the evolution of the pandemic, and feedback on how current public health interventions are performing, in order to begin to restore broad-based trust in the scientific method.

## Data Availability

All data are fully available without restriction and referenced as publicly available data.

## ACKNOWLEDGEMENT

We would like to acknowledge Dr. Alan Maloney for his helpful edits and suggestions in reviewing the drafts of this manuscript.

